# Development and validation of cost-effective one-step multiplex RT-PCR assay for detecting the SARS-CoV-2 infection using SYBR Green melting curve analysis

**DOI:** 10.1101/2021.05.06.21256629

**Authors:** Shovon Lal Sarkar, A. S. M. Rubayet Ul Alam, Prosanto Kumar Das, Md. Hasan Ali Pramanik, Hassan M. Al-Emran, Iqbal Kabir Jahid, M. Anwar Hossain

## Abstract

TaqMan probe-based expensive commercial real-time (RT) PCR kits are being used in COVID-19 diagnosis. The unprecedented scale of SARS-CoV-2 infections has urgently needed to meet the challenge of testing more persons at a reasonable cost. This study developed a rapid, simple, and cost-effective alternative diagnostic method based on melting curve analysis of SYBR green multiplex assay with a host-specific internal control. A total of 90 randomly selected samples were used for comparing the assay with an available commercial kit to analyse the variation and validity of this in-house developed method. Our customized designed primers specifically detected the virus as similar to commercial kit manufactured by Sansure Biotech Inc. We optimized separately the N, E, S, and RdRp genes by SYBR Green RT-PCR method based on melting curve analysis. Afterwards, a multiplex COVID-19 diagnosis method targeting N and E genes of the virus along with the β-actin gene of the host as an internal control has been established. The total run-time of our proposed method was less than 90 minutes. The cost of each sample processing was less than $2. Overall, this one-step and one-tube method can revolutionize the COVID-19 diagnosis in developing countries.

## Introduction

The outbreak of pneumonia like symptoms among the people in Wuhan, China caused by unknown etiology made a global concern since in December 2019.^1^ World Health Organization (WHO) declared the disease as a pandemic coronavirus disease 2019 (COVID-19) caused by Severe acute respiratory syndrome coronavirus 2 (SARS-CoV-2) in March 2020.^2^ The continuous spread of COVID-19 pandemic demands a low-cost diagnosis to stop person-to-person viral transmission. Commercial or in-house RT-PCR based on TaqMan chemistry, the gold standard for detecting SARS-CoV-2 for its higher specificity and correlating the viral load with the cycle threshold value, is the only reliable way for detecting SARS-CoV-2^3–5^. Nonetheless, SYBR Green based RT-PCR could be used as a better alternative for its low cost wherein specific fluorescent probes are not required^6^. Several researches have already been performed for establishing SYBR Green based detection method as a cheaper substitute to detect SARS-CoV-2 instead of TaqMan based detection method^6–13^. However, none of them included internal control for host-specific amplification in the multiplex reaction.

The aim of this study is to develop and validate an easy and inexpensive SYBR Green based method to detect SARS-CoV-2 with four primer sets against N, E, RdRp, and S genes including internal control. We compared our study with one TaqMan-based one-step real time PCR Kit provided by the Government of Bangladesh in almost all COVID-19 testing laboratories. Since this study based on SYBR green mimics TaqMan-based one-step qRT-PCR, we performed the analytical sensitivity comparison with clinical samples collected from COVID-19 patients.

## Results

### Primer Sequence Validation

The oligo-analyzer tool showed that no stable secondary structure, no hairpins, no homodimers, and no cross-dimer were formed in the primer set sequences. The result of the Primer-BLAST tool from the NCBI showed that all the genes’ primer set only matched with the expected target size of SARS-CoV-2 virus genome (reference strain Wuhan-Hu-1), indicating that the performance of the SYBR Green would be sensitive enough for SARS-CoV-2 detection. Searching against the human genome and other pathogens showed no similarity, bolstering that the viral genome segments alone would be amplified (supplementary Table S1). We found no mutation that engendered the new variants within the primer binding region of the viral genome and mutations with low frequency were identified within the last five bases of spike forward and reverse primers, while RdRp, N, and E targeted primers were void of end site mutations. The result of the searches was represented in the supplementary Table S1.

### Development of Singleplex SYBR Green assay

Using JUST_N1, JUST_E1, JUST_S1, JUST_RdRp1, beta-actin and GAPDH primer sets, a singleplex assay was initially performed for four clinical samples (3 positive and 1 negative). Specific desired band for each gene target was identified in the gel electrophoresis as mentioned in the table 1 (supplementary figure 1). In this assay, the amplicon of JUST_N1, JUST_E1, JUST_S1 and JUST_RdRp1 primers produced a specific Tm peak at 82.32±0.17°C, 79.40±0.31°C, 76.52±0.17°C and 77.57±0.17°C, respectively in the melt curve for positive samples. However, a clearly distinct dissociation curve was generated for negative samples that varied with the melt curve as identified for the positive samples, suggesting non-specific signals or the formation of primer dimers in the melt curve peaks for the negative samples. The amplicon of housekeeping gene β-actin and GAPDH produced a specific Tm peak at 87.59±0.18°C and 82.95±0.036°C for both positive and negative samples (Figure 2).

**Table 1:**
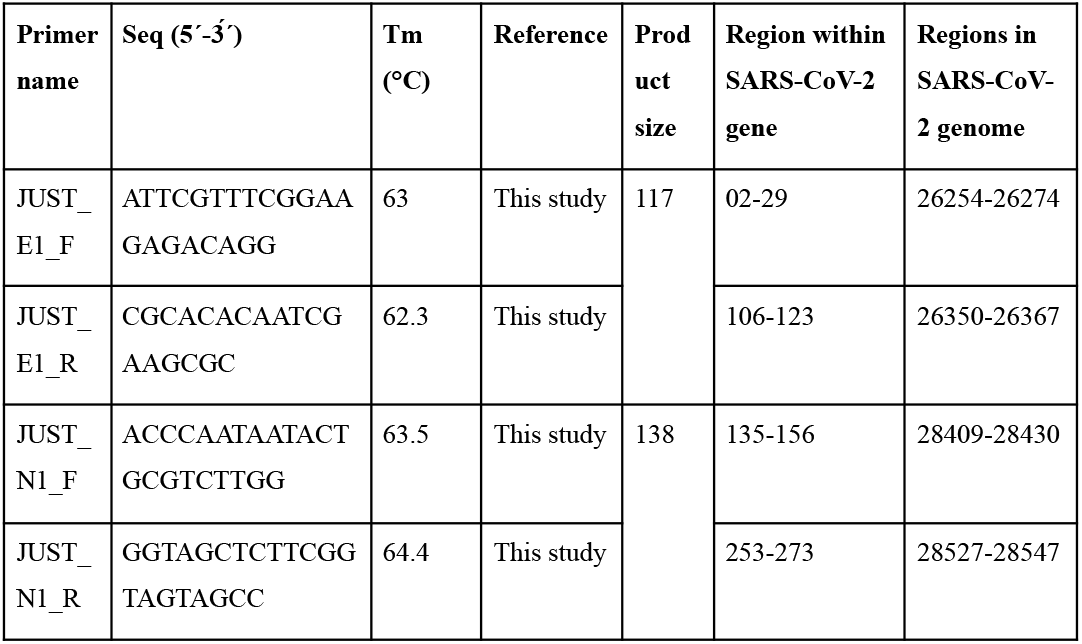
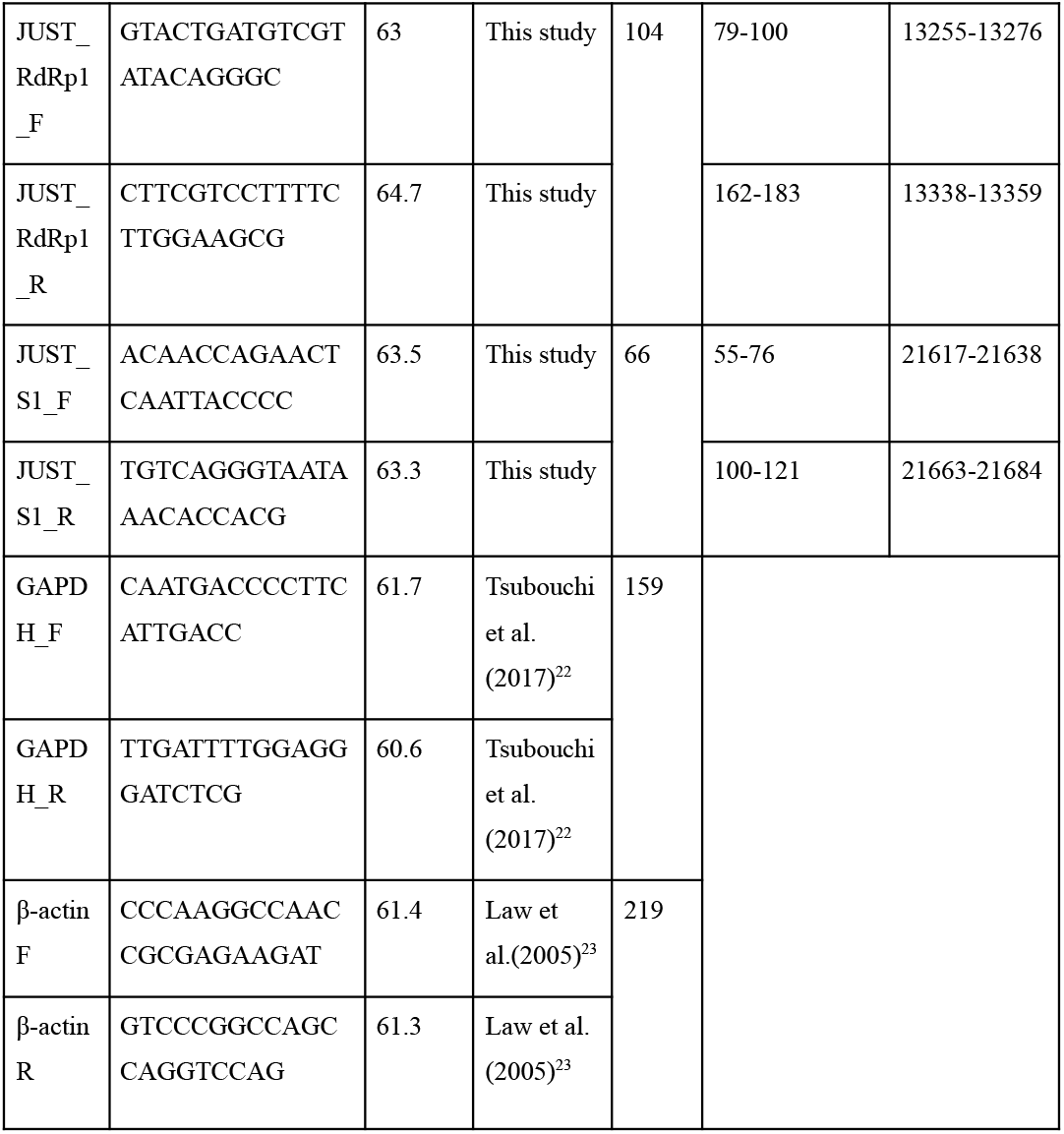

**Figure 1:**
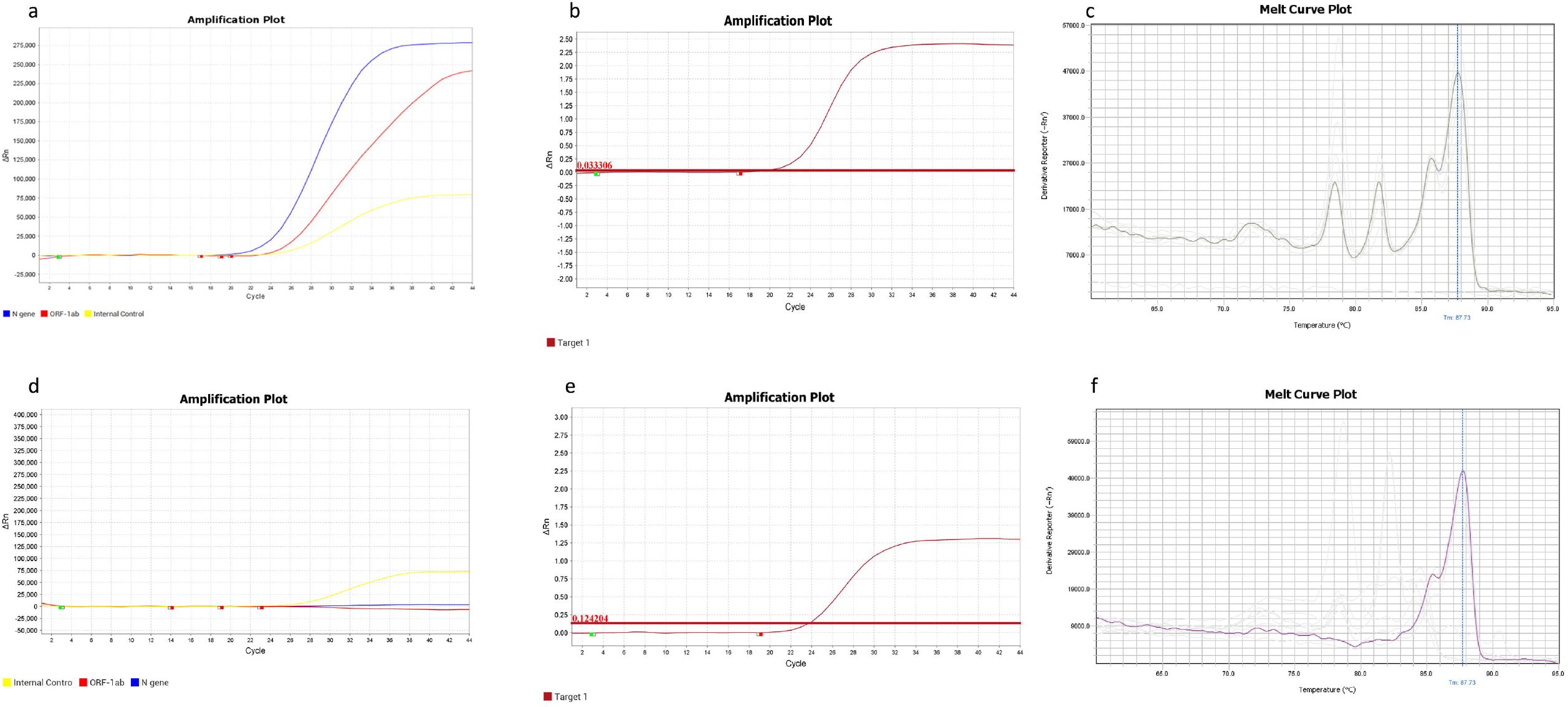
a) Positive result for TaqMan Based method; b) Positive result for N, E and β-Actin genes in SYBR Green Based Method (linear View) ; c) Positive result for N, E and β-Actin genes in SYBR Green Based Method (Melt curve plot) d) Negative result for TaqMan Based method (only Internal control peak); e) Negative result for SYBR Green Based Method (linear View); f) e) Negative result for SYBR Green Based Method (Melt Curve Plot);

**Figure 2:**
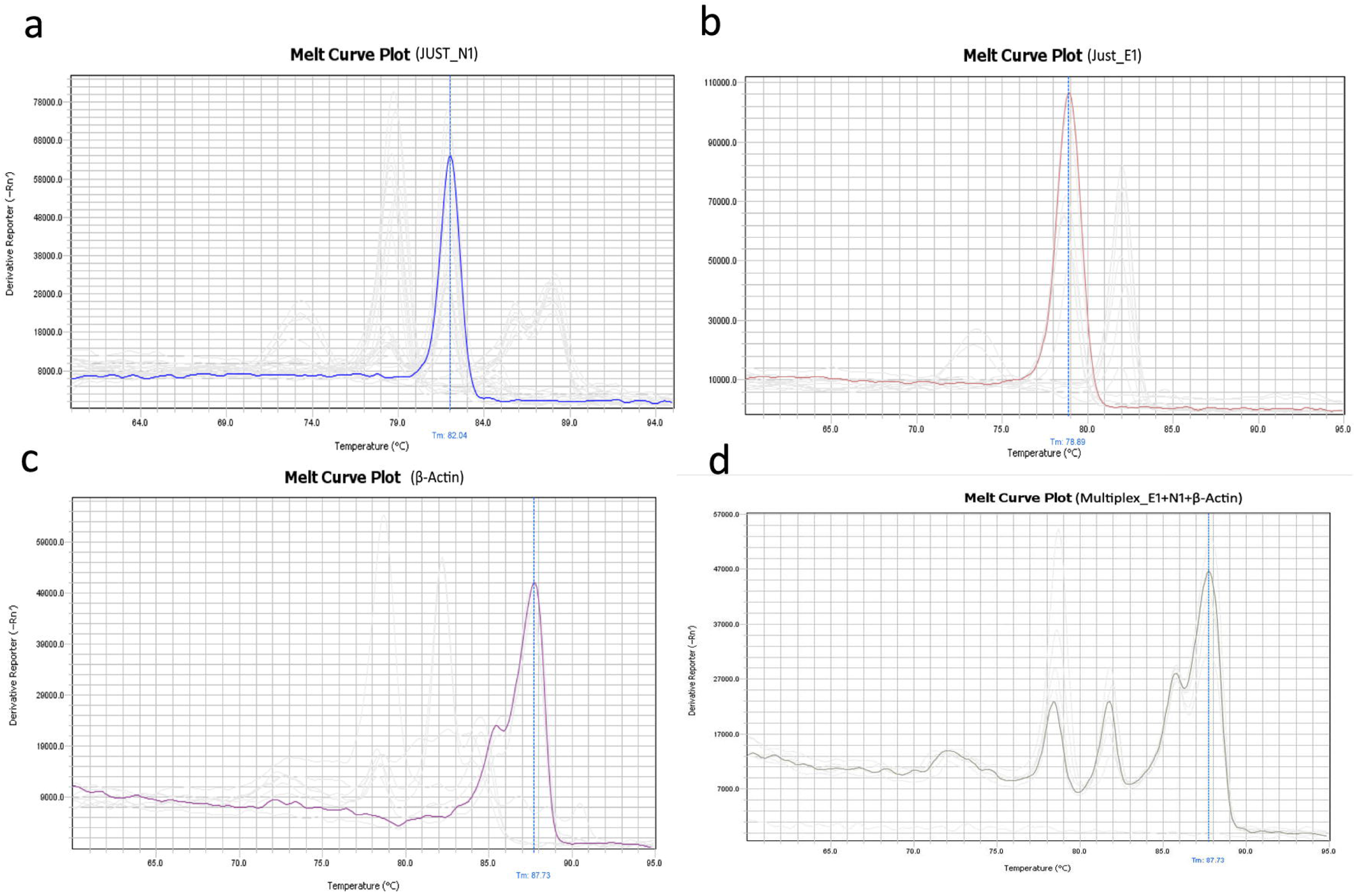
Melting curve plot of singleplex and multiplex assay. a) Melting curve of N gene of Covid-19 Positive sample; b) Melting curve of E gene of Covid-19 Positive sample; c) Melting curve of Housekeeping gene β-Actin; Melting Curve of Multiplex E+N+ β-Actin genes of Covid-19 Positive Sample.

### Optimization of multiplex SYBR green assay

Using the above mentioned primer sets in nine different combinations: JUST_N1+JUST_E1; JUST_N1+JUST_S1; JUST_N1+JUST_RdRp1; JUST_E1+JUST_S1; JUST_E1+JUST_RdRp1; JUST_S1+JUST_RdRp1; JUST_N1+GAPDH; JUST_E1+GAPDH and JUST_S1+GAPDH, we conducted duplex assay and in five different combinations: JUST_N1+JUST_E1+JUST_S1; JUST_N1+JUST_RdRp1+JUST_S1; JUST_N1+JUST_RdRp1+JUST_E1; JUST_N1+JUST_E1+β-actin and JUST_N1+JUST_E1+GAPDH used to conduct triplex assay for two positive clinical samples and no template control (NTC). Quadruplex assay was also performed using primer sets in combination of JUST_N1+JUST_E1+JUST_S1+JUST_RdRp1 for those positive samples.

In duplex assay, the amplicon of JUST_N1+JUST_E1; JUST_N1+JUST_S1; JUST_N1+JUST_RdRp1; JUST_E1+JUST_RdRp1; JUST_N1+GAPDH; JUST_E1+GAPDH, and JUST_S1+GAPDH produced specific Tm peak at (81.58±0.97°C and 78.30±0.85°C); (81.97±0.43°C and 75.59±0.27°C); (82.09±0.18°C and 76.97±0.15°C); (78.90°C and 76.50°C); (82.26°C and 83.90°C); (78.89°C and 82.42°C) and (75.82°C and 82.42°C) respectively. But the amplicon of JUST_E1+JUST_S1 and JUST_S1+JUST_RdRp1 produced only single specific Tm peak at 78.95°C and 77.01°C respectively, which posed that the amplicon of JUST_S1 was not properly amplified.

In triplex assay, the amplicon of JUST_N1+JUST_E1+JUST_S1; JUST_N1+JUST_RdRp1+JUST_S1 and JUST_N1+JUST_RdRp1+JUST_E1 produced two specific Tm peaks at (82.42°C and 76.02°C); (82.27°C and 77.23°C); and (82.27°C and 79.07°C), respectively. Three specific amplicons were produced properly in the triplex assay of JUST_N1+JUST_E1+ β-actin and generated Tm peak at 81.82°C, 78.49°C and (85.78°C and 87.73°C – a signature Tm peak for β-actin). On the other hand, the amplicon of housekeeping gene GAPDH in combination with JUST_N1 and JUST_E1 in triplex assay was not amplified properly and produced two specific Tm peaks at 82.20°C and 78.43°C, respectively.

Finally, in quadruplex assay of JUST_N1+JUST_E1+JUST_S1+JUST_RdRp1, only two amplicons were amplified and produced two specific Tm peak at 82.12°C and 77.54°C. Since the amplicon size of JUST_S1 is small (66 bp), thus could not be differentiated from primer-dimer (as it produced Tm peak at lower temperature and similar Tm peak also appeared for primer-dimer or due to non-specific amplified or fragmented products). We also eliminated JUST-RdRp1 from multiplex assay for small product size (104 bp) and less accurate result in the melting curve with other gene targeting PCR and also eliminated housekeeping gene target in primer GAPDH as it produced closer Tm peak with JUST_N1. The result was summarized in supplementary table 3.

### Validation of in-house established Multiplex SYBR Green Assay with clinical samples

For initially establishing optimal SYBR green based protocol, we used three positive samples and one negative sample as previously confirmed by probe based commercial kit where singleplex qRT-PCR was performed using different sets of primer (JUST_N1, JUST_E1, JUST_S1 and JUST_RdRp1) at different concentration (50-500 nM) and different annealing temperature (58°C-68°C). In our study, the optimal condition of annealing temperature was set at 62°C and 200 nM primer concentration. We also lessened the annealing and extension time (combinedly 25 seconds instead of 1 minute as per standard protocol) to reduce the primer dimer formation and getting a higher quantity of target amplicons.

In this study, the amplicon of JUST_N1 primer and JUST_E1 primer produced a specific Tm peak at 81.8 ± 0.40°C and 78.4 ± 0.33°C in the melt curve for positive samples. For internal control, β-actin primer amplified product generated specific Tm peaks in two positions at around 85.8 ± 0.3°C and 87.8 ± 0.3°C, that is a signature peak (Figure 2). We compiled the singleplex into a multiplex PCR assay based on melting curve and with some modifications to simultaneously detect multiple genes of SARS CoV-2. However, a clearly distinct dissociation curve was generated for negative samples that varied with the melt curve as identified for the positive samples, suggesting non-specific signals or the formation of primer dimers in the melt curve peaks for the negative samples. This is the crucial parameter in the analysis of specificity of curves for the SYBR Green methodology. In the standard curve, the Ct value for JUST_N1 were 19.42, 23.66, 25.64, 27.18, and 27.62 whereas for JUST_E1 were 19.92, 23.20, 26.32, 27.65, and 28.24 for 10^−1^, 10^−2^, 10^−3^, 10^−4^, and 10^−5^ dilution of template RNA (Figure 3). The R^2^ value of 0.9386 and 0.9172 for the N and E gene specific qPCR showed a good correlation between viral copy numbers and Ct values (Figure 4).

**Figure 3:**
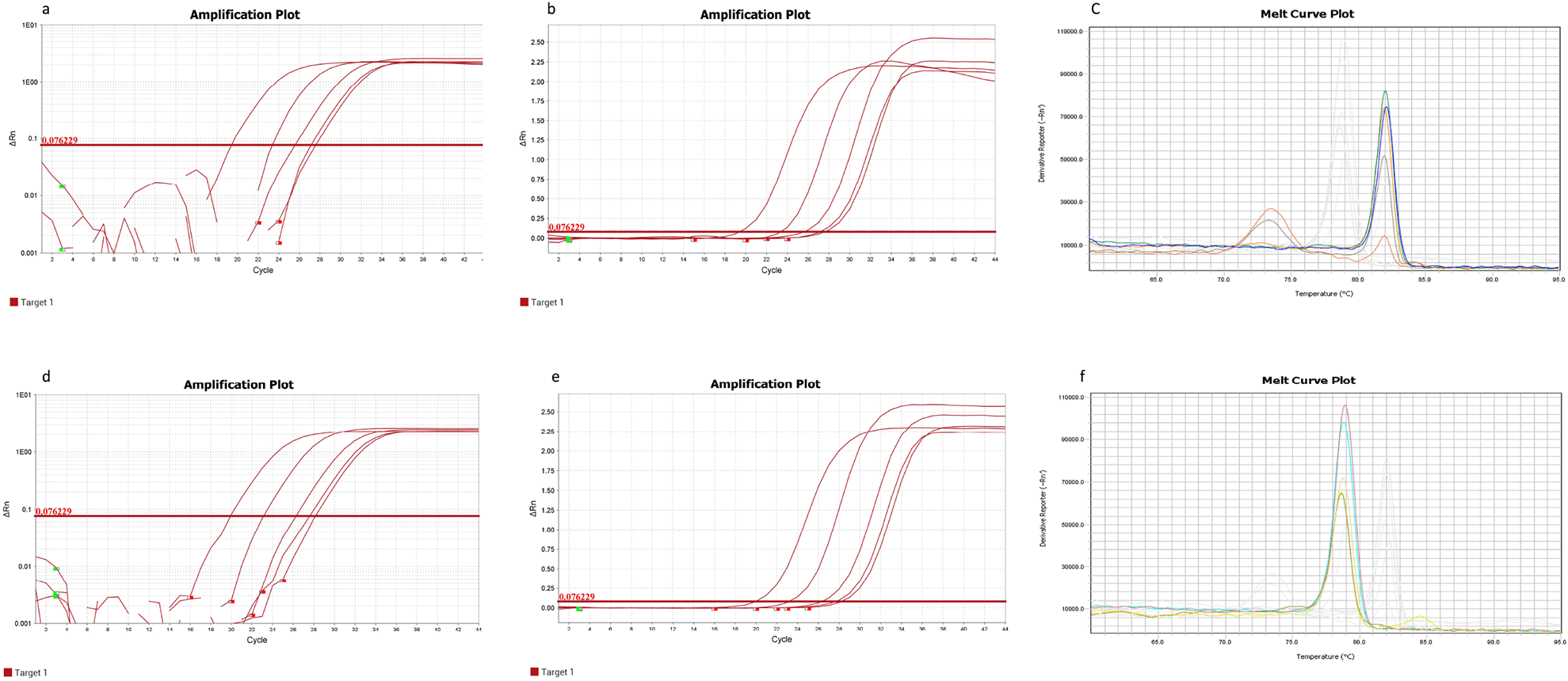
Standard Curve of Just N1 and Just E1 genes. a) Amplification plot of JUST N1 gene (Logarithmic View) in different dilutions (10^−1^ to 10^−5^); b) Amplification plot of JUST N1 gene (Linear view) in different dilutions (10^−1^ to 10^−5^) c) Melting curve of JUST N1 genes in different dilutions; d) Amplification plot of JUST E1 gene (Logarithmic View) in different dilutions (10^−1^ to 10^−5^); e) Amplification plot of JUST E1 gene (Linear view) in different dilutions (10^−1^ to 10^−5^) f) Melting curve of JUST E1 genes in different dilutions;

**Figure 4:**
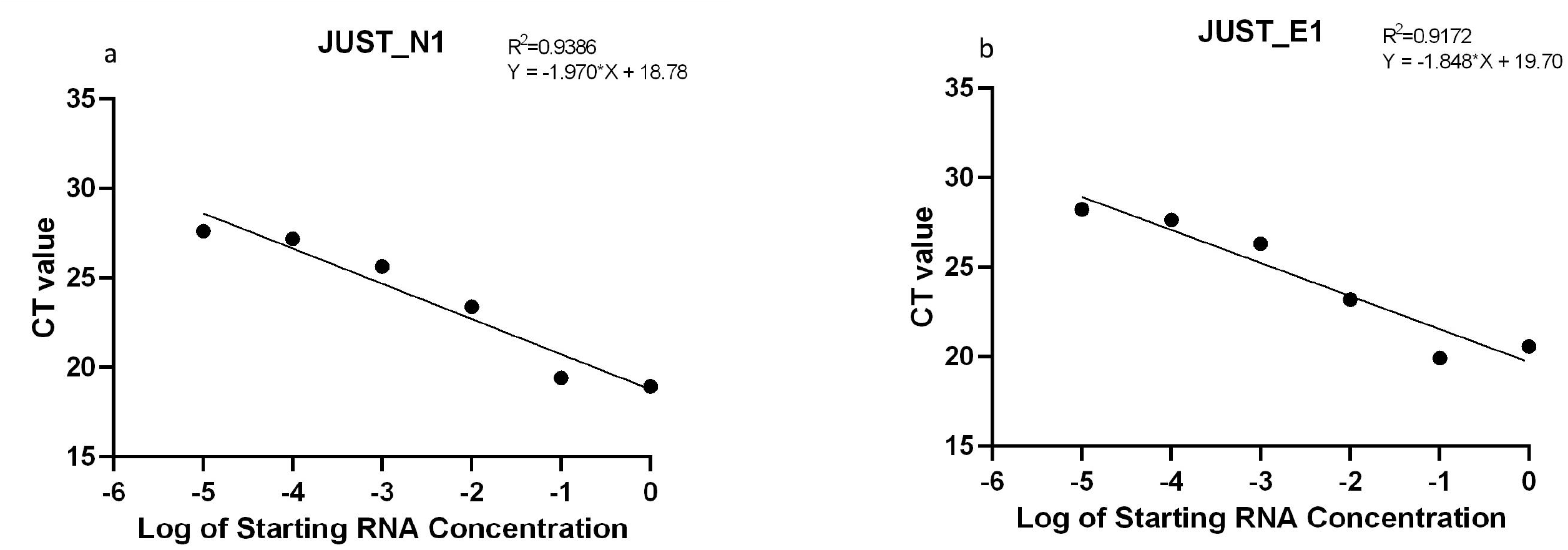
Calibration of qPCR based SYBR green assay. a) Serially diluted targeted RNA of N gene and b) Serially diluted targeted RNA of E gene. The coefficient of determination (R^2^) and linear regression curve (y) were determined.

### Validation of SYBR Green assay against Probe-based Method

In the optimization step, we assayed 90 samples from COVID-19 cases with commercially available TaqMan based kit and our SYBR Green-based One-Step RT-qPCR protocol with two primer sets for SARS-CoV-2 nucleocapsid gene (JUST_N1), envelope gene (JUST_E1) and a primer set for housekeeping gene (β_actin). In both qPCR methods, 49 samples showed positive and 8 samples were negative in both of the RT-qPCR assays (Table 2 and Figure 2). However, we observed 33 samples with contradictory false positive or false negative results (supplementary Table S2). To validate the results, we performed the targeted PCR amplification of spike protein coding sequence and observed product in gel electropherogram (unpublished data). We found a 33% (9/27) positive and 67% (4/6) negative results that corroborated with our SYBR-green based assay for these conflicting samples. On the other hand, only a 33% (2/6) positive and 38% (10/27) negative results accorded with the TaqMan method. Furthermore, we selected three representative samples (one positive and two contradictory) to perform Sanger sequencing using BigDye™ Terminator v3.1 Cycle Sequencing Kit (ThermoFisher Scientific) to estimate the amplicon validation.

**Table 2:**
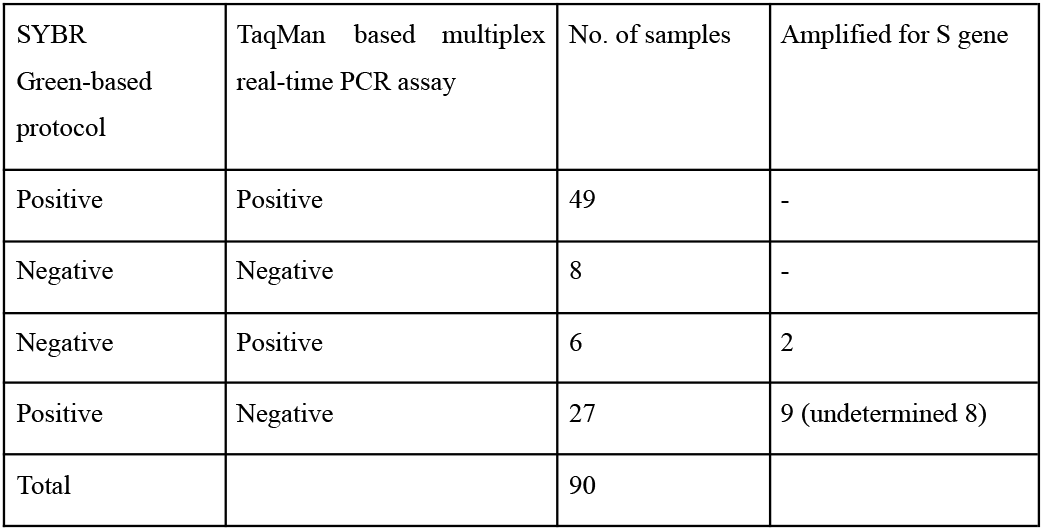

| SYBR Green-based protocol | TaqMan based multiplex real-time PCR assay | No. of samples | Amplified for S gene |
| --- | --- | --- | --- |
| Positive | Positive | 49 | - |
| Negative | Negative | 8 | - |
| Negative | Positive | 6 | 2 |
| Positive | Negative | 27 | 9 (undetermined 8) |
| Total |  | 90 |  |

### Comparison between the Methods

The melting curve peak basically shows the presence of a particular gene in a sample and the derivative reporter is a measurement for the approximate quantity of the present gene. Either N or E gene was detected in 27 out of 49 SYBR positive-Sansure kit positive samples and 17 such cases were present out of 27 SYBR positive-Sansure kit negative samples. In the case of N genes in SYBR positive-Sansure kit positive samples, there were only 10 samples out of 27 where the threshold of derivative reporter went up and beyond 20,000 whereas there were one such sample out of 12 in SYBR positive-Sansure kit negative samples. On the other hand, E gene specific primers could detect 43 and 26 samples in SYBR positive-Sansure kit positive SYBR positive-Sansure negative samples, respectively. The mean value of derivative reporter for SYBR positive-Sansure kit positive was higher by 2,283 with similar standard deviation of ∼5000.

### Sanger Sequencing Results

We selected three representative samples (both methods positive: GC164.02, SYBR positive-Sansure negative: GC164.36, and SYBR negative-Sansure positive: GC165.104) from the 18 amplified spike targeted products among the contradictory resulted samples to perform Sanger sequencing to estimate the amplicon validation and identified matching with the SARS-CoV-2 spike both in BLAST (>99%) and MEGA7 based alignment (supplementary figure S2).

## Discussion

COVID-19 cases are surging worldwide and the developing countries have faced the obstacle to test a large number of samples to control the spread of the disease. Despite a higher price per sample, laboratories and diagnostic centers preferred the probe-based method to detect SARS-CoV-2. A convenient SYBR Green assay as we established here can diagnose with similar efficiency by low cost (supplementary table S3). For diagnostic purposes, it is convenient to mimic the regularly used Taqman based commercial kit assay using a multiplex SYBR Green method so that the regular users will face no hindrance to apply. Generally, SYBR green method has some issues of nonspecific signaling by binding primers to the unwanted regions of the sequence, formation of primer-dimer, and presence of remaining segmented templates. Elimination of this factor by gauging primer sequence and PCR condition would significantly increase the acceptance of this technique as a diagnostic tool and optimization of the method can improve its performance and quality to be comparable to a standard TaqMan assay^6^.

In this study, the designed primers were successfully sifted out the non-specificity issues, and also by troubleshooting the annealing temperature of our primers in different combinations, we could detect the specific genes of only SARS-CoV-2 given that other human coronaviruses might present in the clinical samples. Besides, the primers are targeted solely against the SARS-CoV-2 with the screening out of the mutations present among the GISAID global viral sequences within the last five bases of the 3’-end of the primer. However, we found a partial matching of the RdRp, N, and E primers with the SARS coronavirus Tor2 genome (supplementary table S1), and it is noteworthy that SARS was not detected in human and other animals after 2005^16^. These criteria, together, can give highly specific results for our developed SYBR green assay.

Four customized primer sets against N, E, RdRp and S gene were employed in this method for two reasons. Firstly, the N and S are the most expressed transcript during viral replication ^17^. Secondly, the E and RdRp are the least evolving genes due to minimize effects of the selective pressure ^18^. Since the amplicon size of JUST_S1 is small (66 bp), thus could not be differentiated from primer-dimer and there had also arisen some cases of mutations within the last 5 bases of the S targeting primer region (supplementary Table S1) while carrying out the experiment, hence we eliminated the S targeting primers from further multiplex study. We also eliminated JUST-RdRp1 from multiplex assay for small product size (104 bp) and less accurate result in the melting curve with other gene targeting PCR. We initially optimized the condition for each gene and then targeted multiple genes for conducting multiplex PCR by combining virus-specific primer sets along with an internal control gene of the host. While most of the previous studies were performed based on two step RT-PCR (cDNA preparation and PCR reaction in different tubes), we performed those in a one step RT-qPCR using a single tube (cDNA preparation and PCR in the same tube) that has facilitated the workflow and also reduced the chance of contamination. Gomez et al. (2020) and Dorlass et al. (2020) at first converted extracted RNA into cDNA and then performed the PCR with the cDNA template which is a two-step process and relatively time-consuming^9,12^. However, we performed only one step RNA extraction in a single tube that after adding the qRT-PCR reagents lead to the subsequent RT-qPCR.

Our designed primers were screened against the covariants and clade-featured mutations that independently originated in different parts of the world, now spreading quickly, and importantly, arised after most of the previous SYBR green based methods^19^, thus this assay can effectively detect the variants. Another supremacy of our study is the multiplexing with three genes (N, E, and beta-actin) simultaneously where we included internal control β-actin, which is a host-specific gene. β-actin here was used for the same purpose as in TaqMan assay for confirming the human sample and whether the sampling has been collected accurately from the nasopharyngeal swab. This engendered a unique approach and was not present in any current SYBR-Green based study. When multiple genes are used in the SYBR green method, it is not feasible to predict the presence or absence of any particular gene or primer dimer by specific Ct-value. Melting curve-based detection therefore solved the issue wherein the dissociation point of three genes are different and any flat curve on that melting point predicted the absence of the specific gene in the sample. A slight deviation from the average melting temperature in the dissociation curve for the N (81.8 ± 0.40°C) and E (78.4 ± 0.33°C) genes would mean minimal or no sequence variation among the viruses present in different clinical samples. The difference between the derivative reporters (mean = 2,283) of the SYBR positive-Sansure kit positive and SYBR positive-Sansure kit negative samples was most probably due to the lowest quantity of the virus present in those samples. Even though we used a lower amount of RNA for the assay (upto 7 ul), we derived better results in terms of detecting the virus showing a less value of derivative reporter for those samples (supplementary table S2). Since our target was not to quantify viruses present in a sample and compare the assays based on quantification, we did not proceed further on this issue.

Pereira-Gómez et al. (2021) used primer sets against ORF1b-nsp14 or N gene targets and found that SYBR Green based assays were consistent with the TaqMan probe-based protocol^12^. They also performed melting curve analysis to differentiate primer-dimer from the target product amplification. Park et al. (2020) established a multiplex cost-effective SYBR Green based method using primer sets targeting RdRp, N, E, and S gene while suggesting melting curve analysis for confirming the specific target product amplification. However, their work did not include the internal control specific primers in the multiplex reaction^10^, that we included in this study. Pearson et al. (2020) compared TaqMan and SYBR green based detection method for N and S genes from previously published primers but they did not design any SYBR green based detection method^14^. Erick Gustavo Dorlass et al. (2020) also conducted a SYBR-Green assay for the detection of SARS CoV-2 E gene in clinical samples but the multiple gene detection in SYBR-Green method is absent in their study^9^. Our study thus is superior to other studies since this method generated the similar results as the probe-based assay to separate the disparate COVID-19 positive and negative samples.

Finally, our target was not to quantify rather detect the presence or absence of virus in the clinical samples that we have successfully accomplished here. Nevertheless, this study has some limitations. We could not be consistent with the gold standard outcomes obtained by commercial kits with respect to Ct value measured for each gene. Discrepant results wherein one gene was detected in SYBR green assay were observed in more than half of the samples for both SYBR positive-Sansure kit positive (27/49) and SYBR positive-Sansurel kit negative groups (17/27). Other researchers however reported the similar trend despite with a very small percentage of the samples ^20^. Using a lower amount of RNA template, the less reliable results than TaqMan assay, multiplexing of three genes in SYBR Green, and arises of possible mutations during the study in highly dynamic nucleocapsid protein^20,21^ might reduce the chance to amplify N gene specific region. Moreover, the sensitivity and the specificity were not completely determined due to not validating all the studied samples, though an identical sensitivity and specificity was determined while comparing the 33 deviated samples between SYBR Green and TaqMan assay (Table 2). The reasons behind a low R^2^ value (0.938 for JUST_N1 and 0.917 for JUST_E1) might be the presence of non-specific RNA due to a quick RNA extraction system, possible presence of polymerase inhibitors, and formation of primer-dimer. Another subtle limitation of our technology is that only expert personnel will be able to analyze the results since it is based on melting curve analysis. SYBR Green technique was suitable for detecting SARS-CoV-2 in clinical samples; and our user friendly and affordable protocol detected clinical SARS-CoV-2 as efficiently as a standard costly Taqman protocol. Since this SYBR green based method of detection SARS-CoV-2 performed well after optimization and the results can be easily predicted by melting curve analysis, we recommend this method as an easier, cheaper, and reliable alternate to the costly probe-based method to increase the testing capacity for low and middle income countries where reagent supply is limited and high testing capacity is desired.

## Methods

### Clinical sample selection and ethical consideration

The sample size was calculated based on the prevalence of positive samples using the following equation

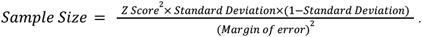

Randomly (random number generator using Microsoft Excel inc.) selected 6 positive and 4 negative samples were selected from left-over samples once in a week and tested by our proposed method besides the routine TaqMan based method. In 9 weeks from 10 October 2020 to 30 November 2020, we collected a total sample size of 3836 where 560 samples were positive. Using the above equation with a 10% margin of error and 95% confidence level for the samples and finally a total of 90 nasopharyngeal swab samples were selected for both methods. All patients’ samples were selected from the continuous surveillance system screening for COVID-19 at the Genome Center, Jashore University of Science and Technology (JUST) authorized by the Directorate General of Health Services (DGHS), Bangladesh. The study was approved by the ethical review committee (ERC) of Jashore University of Science and Technology, Bangladesh (Reference: ERC/FBS/JUST/2020-45, Date: 06/10/2020).

### Primer designing for SARS-CoV-2 strains

We designed our primers by targeting N, E, S, and RdRp protein coding nucleotide sequences based on the aligned sequence data of all circulating SARS-CoV-2s available in GISAID. We used Nextstrain (https://covariants.org/variants) and National Genomics Data Center (https://bigd.big.ac.cn/ncov/variation/annotation) for checking whether the mutation sites fall within the primer binding region or not. We also checked the primers against the genome of *Homo sapiens*, six common human coronaviruses HCoV-OC43, HCoV-229E, HCoV-NL63, HCoV-HKU1, MERS-CoV, and SARS-CoV, and finally main respiratory and opportunistic viruses and pathogens in PrimerBLAST (supplementary table S1). As the internal control, we primarily targeted three house-keeping genes of the human genome, GAPDH and beta-actin. We designed the SARS-CoV-2 specific primers in a way to maintain the annealing temperature at ∼60ºC for efficient qPCR detection and then used oligo-analyzer tool to check the primers for stem-loops and highly energetic dimer formation (< -9 Kcal/mol). The amplicons’ size was distinct so that we could identify separate products in the melting curve (**table 1**).

### Crude RNA Extraction from the Samples

The viral RNA was extracted using QuickExtract™ RNA Extraction Kit (Lucigen, USA) according to the manufacturer’s instruction (cat number: QER090150). Briefly, a particular volume of the samples (5-100 µl) VTM was separated from patients swab sample inside Biosafety Cabinet (Class II) and mixed with an equal volume of ice-cold QuickExtract RNA Extraction Solution (5-100 µl) in Eppendorf tube and vortex-mixed for 1 minute, followed by immediate transfer into ice.

### Commercial Fluorescence-based RT-PCR

In TaqMan probe-based RT-qPCR method, commercially available SARS-CoV-2 nucleic acid detection kits (Sansure Biotech, China) were used to compare with in house SYBR Green kit. The kit contains SARS-CoV-2 ORF1ab and N genes, and as internal control, human IRC genes (i.e. Rnase P). The reaction conditions and procedures were applied according to protocol described elsewhere and all reactions were performed in duplicate to confirm reproducibility. The reaction systems and procedures were carried out according to the instructions of the kits (http://eng.sansure.com.cn/index.php?g=&m=article&a=index&id=81). In brief, 13μl 2019-nCoV-PCR Mix was mixed with 2μl 2019-nCoV-PCR-Enzyme Mix and added to 10μl template RNA. The RT-qPCR conditions were set according to the manufacturer’s instructions as follows: reverse transcription at 50°C for 30 minutes, cDNA pre denaturation at 95°C for 1 minute, then denaturation at 95°C for 15 seconds and annealing, extension and fluorescence collection at 60°C for 30 seconds for 45 cycles before cooling the device at 25°C for 10 seconds in Real Time PCR machine (QuantStudio 3.0, Applied Biosystem). For detection, FAM, ROX, and VIC were used to detect ORF1ab, N gene, and internal control, respectively. Supplied Positive control and supplied negative control with kit as well as nuclease-free water was included in every qPCR run as a positive control, kit negative control and reaction negative control, respectively. Sigmoid curve for either or both of the ORF1b-nsp14 or N gene with a CT value of ≤ 36 was interpreted as positive. CT values between 37 and 39 were repeated and above those (≥40) were considered negative in the prevalence study.

### Optimization of Singleplex RT-qPCR

To detect SARS-CoV-2 target genes, melting curve-based RT-PCR was performed using SYBR Green fluorescent dye, which binds double-stranded DNA by intercalating between the DNA bases. All RT‐qPCRs were performed on Applied Biosystems QuantStudio3 Real‐Time PCR Systems and Design and Analysis Software v1.5.1, using 0.2ml MicroAmp™ Optical 96-Well Reaction Plate (Cat. No. N8010560) and MicroAmp™ Optical Adhesive Film (Cat. No. 4311971). Initially, commercial kit based repeatedly confirmed positive and negative samples were considered to check the accuracy and efficiency of each primer set. We verified this through one step RT-qPCR amplification (QuantStudio 3.0: Applied Bioscience) of the amplicons to optimize the PCR conditions and primer set concentration. The target SARS-CoV-2 genes included N1, S1, E1, and RdRp1. In addition, either GAPDH, or β-actin of humans, which is a housekeeping gene, was used as an internal positive control. For each reaction, 5μl of extracted RNA template was used for the SARS-CoV-2 specific target primer sets and human internal positive control primer set. Gradient RT-qPCR was performed with an increasing annealing temperature (Tm) from 60°C to 70°C based on the melting temperature of each primer set. Different concentrations of primer were used for optimizing concentration of the primer sets.

The basic RT-qPCR conditions were set according to the Luna^®^ Universal SYBR green One-Step RT-qPCR Kit (New England Biolabs Inc, MA). Detailed protocol is provided in the manufacturer’s recommended procedures (https://international.neb.com/products/e3005-luna-universal-one-step-rt-qpcr-kit#Product%2 0Information). We mixed 10μl master mix (2X) with 1μl enzyme mix (20X), 200 nM forward primer, and 250 nM reverse primer for each gene in single primer set. The PCR conditions were set at 58°C incubation for 12 minutes for reverse transcription followed by the initial denaturation at 95°C for 3 minutes, then cycle denaturation at 95°C for 15 seconds and an extension at 62°C for 25 seconds for 45 cycles. For the melt curve analysis, we set 95°C for 15 seconds, then 60°C for 1 minutes followed by 95°C for 15 seconds. The ramp rate of last transformation of 60°C to 95°C was set at 0.05°C/seconds. For passive reference, ROX was used as the passive reference and SYBR dye was used to check the fluorescence in the Real Time PCR machine (QuantStudio 3.0, Applied Biosystem). Low Ct valued samples (<20 as detected in TaqMan method) and nuclease-free water were included in every qPCR run as a positive control and negative control, respectively.

### Optimization of Multiplex RT-qPCR

For the multiplexing, two sets of SARS CoV-2 specific primers, JUST_N1 and JUST_E1, as well as, β-actin (human control) primer set were used since the amplified products had distinct melting curve peaks — that eased the identification of different products. The primer concentration was optimized for multiplex RT-qPCR as follows: JUST_N1 forward 300 nM, JUST_N1 reverse 400 nM, JUST_E1 forward 150 nM, JUST_E1 reverse 175 nM, β-actin forward 150 nM, and β-actin reverse 200 nM. 10μl master mix (2X) with 1μl enzyme mix (20X) of Luna^®^ Universal One-Step RT-qPCR Kit (New England Bioscience) with 5μl of template RNA was used under the PCR conditions as performed for the singleplex one. We set the criteria to determine the positive and negative results: presence of any peak for either N or E genes in the melting curve. The results were not interpreted based on the derivative reporter value of the melting curve for each gene since a peak with low value but good shape was considered in this case. The overall protocol of the assay has been presented within a single kit-based manual style (Figure 5).

**Figure 5:**
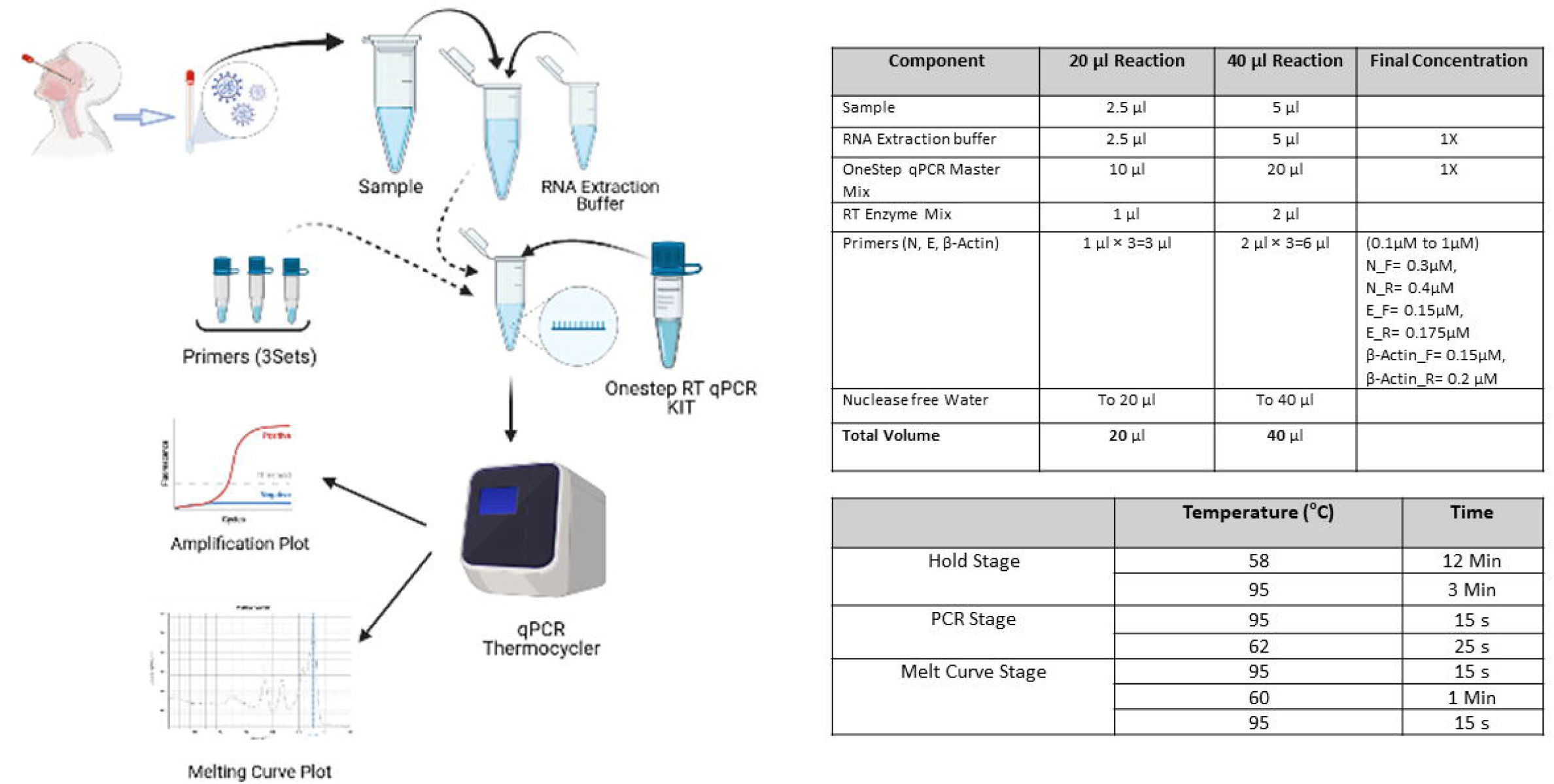
Standard working Procedure

### Generation of Standard Curve and Melting Curve Analysis

For preparing the standard curve, template RNA was diluted 10-fold for 5 times (10^−1^ to 10^−5^). The diluted template RNA was used to conduct single-plex RT-qPCR assay for both JUST_N1 and JUST_E1. The derivative reporter values of the melting curve as measured for the N and E genes were analyzed to compare between the SYBR positive-Sansure positive and SYBR positive-Sansure negative groups. We targeted to set a threshold based on the derivative reporter value from the melting curve.

### Validation by Gel Electrophoresis and Targeted Amplification

We performed gel electrophoresis in 3% agarose gel having ethidium bromide at 60V and 100mA for 100 minutes to ensure amplification of the correct RT-qPCR products and analyzed in an automated Gel Doc Imager (Molecular Imager® Gel Doc™ XR+ System with Image Lab™ Software by Bio-Rad (Catalog # 170-8195) and the software Image Lab™ Software version 5.2.1. To validate the ambiguous results between the SYBR-Green and commercial RT-qPCR assay, the in-house established primer set targeting the spike gene for amplification established by Islam et al. was used and run in 1.5% agarose gel at 80V and 200mA for 40 minutes to ensure amplification^15^.

### PCR Product Purification and Sanger Sequencing

For purifying the desired PCR product from the agarose gel, we used Wizard® SV gel and PCR Clean-Up system. The four representative amplicons were then subjected to Sanger sequencing with BigDye™ Terminator v3.1 Cycle Sequencing Kit (Thermo Fisher Scientific) in Applied Biosystems SeqStudio genetic analyzer as per the optimized protocol of Islam et al. (2021). The ab1 files from the Sanger sequencing were analyzed using the Sequencing Analysis Software V6.0 (Thermofisher, USA). NCBI BLAST was performed initially and the alignment to SARS-CoV-2 spike gene was also checked in MEGA7 (https://www.megasoftware.net/).

## Data Availability

We acknowledge GISAID for sharing the sequence data and IDT for giving the opportunity to use the tools for validating primers in silico. We also acknowledge the Ministry of Health and Family Welfare, Bangladesh, for giving us permission for the SARS‐CoV‐2 diagnosis.

## ACKNOWLEDGMENTS

We acknowledge GISAID for sharing the sequence data and IDT for giving the opportunity to use the tools for validating primers in silico. We also acknowledge the Ministry of Health and Family Welfare, Bangladesh, for giving us permission for the SARS‐CoV‐2 diagnosis. The present study was funded by the Jashore University of Science and Technology Research Grant (#FoBST‐06) supported by the University Grant Commission, Bangladesh.

## Authors’ Contributions

P.K.D. and M.H.A.P. performed the assay in laboratory experiment. S.L.S. and A.S.M.R.U. designed the experiment, planned the troubleshooting, and wrote the the main manuscript. A.S.M.R.U. designed the primers. H.M.A. planned the sampling portion and advised in established the protocol in laboratory. I.K.J. was involved in the whole working procedure and also wrote the manuscript. M. H.A. supported the workflow and critically reviewed the manuscript. All authors perused the manuscript.

