## supplementary for "Development and validation of cost-effective one-step multiplex RT-PCR assay for detecting the SARS-CoV-2 infection using SYBR Green melting curve analysis"

**Supplementary table 1:**

| **Primer name** | **Mutation in 3' end** | **Speficity of Primer(Homo Sapiens, other organisms*)** | **https://covariants.org/variants** | **https://bigd.big.ac.cn/ncov/variation/annotation** |
| --- | --- | --- | --- | --- |
| JUST_E1_F | No | No target templates were found in selected database: Refseq mRNA (Organism limited to Homo sapiens) | No Mutation | 26270, 26256, 26262, 26257 |
| JUST_E1_R | No | Target template found with SARS-CoV-2 isolate Wuhan-Hu-1 and SARS coronavirus Tor2, complete genome | No Mutation | None |
| JUST_N1_F | No | No target templates were found in selected database: Refseq mRNA (Organism limited to Homo sapiens) | No Mutation | 29422, 29427, |
| JUST_N1_R | No | Target template found with SARS-CoV-2 isolate Wuhan-Hu-1 and SARS coronavirus Tor2, complete genome | No Mutation | 28541, |
| JUST_RdRp1_F | No | No target templates were found in selected database: Refseq mRNA (Organism limited to Homo sapiens) | No mutation | None |
| JUST_RdRp1_R | No | Target template found with SARS-CoV-2 isolate Wuhan-Hu-1 and SARS coronavirus Tor2, complete genome | No mutation | None |
| JUST_S1_F | No | No target templates were found in selected database: Refseq mRNA (Organism limited to Homo sapiens) | No mutation | 21637, 21638, 21621, 21624 |
| JUST_S1_R | No | Target template found with SARS-CoV-2 isolate Wuhan-Hu-1 | No mutation | 23664 |
|  |  | *Organisms investigated (Taxonomy ID): HCoV-HKU1 (290028); HCoV-OC43 (31631); HCoV-NL63 (277944); HCoV-229E (11137); MERS-CoV (1335626); H1N1 (114727); H3N2 (119210); Influenza (untyped) (11309); H5N1 (102793); H7N9 (333278); Influenza B (11520); Respiratory Syncytial Virus (11250); Parainfluenza 1 virus (12730); Parainfluenza 2 virus (1979160); Parainfluenza 3 virus (11216); Parainfluenza 4 virus A (11224); Parainfluenza 4 virus B (11226); Human Metapneumovirus (162145); Human Bocavirus (329641); Mycoplasma spp. (2093). Rhinovirus/Enterovirus (12059); SARS-CoV |  |  |

**Supplementary Table 02**

| Sl No. | Lab ID | SYBR Green Based Result | | | | | | | | | | Commercially available Kit (Sansure) Based Result | | | |  |
| --- | --- | --- | --- | --- | --- | --- | --- | --- | --- | --- | --- | --- | --- | --- | --- | --- |
|  |  | Ct Value | JUST_N1 | | JUST_E1 | | β-actin | | | | Comment | Ct Value for N gene | Ct Value for ORF-1ab | Ct Value for Internal Control | Comment |  |
|  |  |  | Tm (°C) | Derivative Reporter | Tm (°C) | Derivative Reporter | Tm (°C) | Derivative Reporter | Tm (°C) | Derivative Reporter |  |  |  |  |  |  |
|  |  |  |  |  |  | **All Positive** |  |  |  |  |  |  |  |  |  |  |
| 1 | Int 731 | 25.142 | 82.228 | 54,963.891 | 78.678 | 11,980.670 | 85.985 | 18,059.166 | 87.940 | 21,272.609 | Positive | 24.769 | 26.097 | 26.641 | Positive |  |
| 2 | 157.62 | 25.902 | 82.074 | 43,878.445 |  |  | 85.882 | 23,577.688 | 87.785 | 28,807.961 | Positive | 26.505 | 26.948 | 28.712 | Positive |  |
| 3 | 157.02 | 25.583 | 81.714 | 18,466.676 |  |  |  |  | 87.425 | 21,165.930 | Positive | 28.281 | 29.516 | 25.446 | Positive |  |
| 4 | 158.01 | 18.143 | 82.023 | 61,780.219 | 78.369 | 15,874.382 | 85.727 | 20,245.354 | 87.888 | 30,311.070 | Positive | 19.629 | 21.285 | 27.441 | Positive |  |
| 5 | 157.42 | 26.817 | 81.920 | 40,864.320 |  |  | 85.727 | 19,907.092 | 87.785 | 19,104.541 | Positive | 29.846 | 30.760 | 28.119 | Positive |  |
| 6 | 158.19 | 24.192 | 81.662 | 14,340.313 | 77.186 | 13,671.268 | 85.521 | 19,507.586 | 87.528 | 24,515.197 | Positive | 32.689 | 35.282 | 26.869 | Positive |  |
| 7 | 157.54 | 21.496 | 81.868 | 68,510.750 |  |  |  |  | 87.631 | 19,082.664 | Positive | 22.676 | 23.342 | 28.758 | Positive |  |
| 8 | 157.55 | 23.431 | 81.765 | 37,118.063 |  |  | 85.573 | 17,410.832 | 87.580 | 24,995.152 | Positive | 27.466 | 29.065 | 26.819 | Positive |  |
| 9 | 157.44 | 29.152 |  |  | 77.622 | 17,526.877 |  |  | 87.565 | 20,616.449 | Positive | 35.564 | Undetermined | 31.931 | Positive |  |
| 10 | Int 725 | 24.527 | 82.129 | 20,525.418 | 78.791 | 19,824.313 | 85.979 | 12,688.072 | 87.981 | 13,201.334 | Positive | 29.458 | 29.611 | 29.745 | Positive |  |
| 11 | 155.10L | 23.981 | 82.198 | 23,276.420 | 78.859 | 26,242.150 | 86.052 | 17,089.092 | 87.799 | 18,954.260 | Positive | 35.003 | 36.309 | 32.035 | Positive |  |
| 12 | 155.08L | 24.276 | 82.077 | 19,984.512 | 78.791 | 23,012.178 | 85.825 | 13,666.186 | 87.622 | 8,185.792 | Positive | 33.826 | 41.076 | 31.227 | Positive |  |
| 13 | 155.16L | 23.953 | 82.044 | 21,010.381 | 78.756 | 26,889.242 | 85.949 | 18,985.578 | 87.799 | 19,066.668 | Positive | 33.854 | 36.073 | 31.113 | Positive |  |
| 14 | 155.18L | 23.855 | 81.872 | 16,114.951 | 78.586 | 19,742.877 | 85.774 | 18,647.953 | 87.725 | 21,276.943 | Positive | 30.092 | 30.651 | 29.300 | Positive |  |
| 15 | 155.17L | 23.993 | 81.993 | 22,560.957 | 78.704 | 27,945.535 | 85.898 | 19,237.627 | 87.696 | 22,045.998 | Positive | 33.767 | 36.086 | 32.471 | Positive |  |
| 16 | 155.24 L | 24.166 | 81.820 | 25,037.160 | 78.535 | 30,310.512 | 85.620 | 22,864.441 | 87.519 | 19,252.742 | Positive | 34.017 | 34.199 | 31.277 | Positive |  |
| 17 | 155.26L | 23.974 | 81.942 | 23,384.619 | 78.602 | 25,100.133 | 85.744 | 15,979.553 | 87.542 | 16,355.396 | Positive | 34.858 | 37.681 | 33.249 | Positive |  |
| 18 | 155.55L | 24.871 | 81.615 | 38,135.703 | 78.381 | 23,627.805 |  |  |  |  | Positive | 30.185 | 30.102 | 31.939 | Positive |  |
| 19 | 155.30L | 23.982 | 81.890 | 17,213.672 | 78.653 | 24,387.582 | 85.847 | 18,282.744 | 87.645 | 24,406.779 | Positive | 34.192 | 36.756 | 30.842 | Positive |  |
| 20 | 117.199L | 22.344 | 81.924 | 46,414.238 | 78.484 | 15,677.862 | 85.775 | 24,702.352 | 87.777 | 30,949.279 | Positive | 24.036 | 23.157 | 25.826 | Positive |  |
| 21 | 116.194L | 28.454 | 81.821 | 23,544.512 | 78.484 | 19,807.328 |  |  | 88.290 | 22,283.141 | Positive | 33.918 | 33.135 | 35.586 | Positive |  |
| 22 | 116.195L | 26.496 | 81.770 | 14,253.395 | 78.279 | 14,237.033 | 85.569 | 25,767.824 | 87.623 | 33,886.613 | Positive | 32.662 | 32.174 | 29.455 | Positive |  |
| 23 | 116.016L | 25.823 |  |  | 78.279 | 12,644.385 | 85.467 | 22,194.375 | 87.418 | 29,343.906 | Positive | 34.719 | 31.612 | 29.502 | Positive |  |
| 24 | 159.03 | 21.444 |  |  | 78.632 | 18,980.840 | 85.869 | 24,304.836 | 87.922 | 42,250.578 | Positive | 25.098 | 31.781 | 24.418 | Positive |  |
| 25 | 159.49 | 24.759 |  |  | 78.529 | 25,857.199 | 85.715 | 23,548.203 | 87.717 | 33,421.344 | Positive | 24.514 | 28.924 | 30.989 | Positive |  |
| 26 | 159.53 | 25.872 |  |  | 78.427 | 17,288.045 | 85.664 | 20,740.063 | 87.563 | 27,611.568 | Positive | 32.231 | 40.326 | 33.956 | Positive |  |
| 27 | 160.05 | 28.184 |  |  | 78.427 | 28,530.355 | 85.459 | 25,730.330 | 87.871 | 26,064.926 | Positive | 25.976 | 28.153 | 28.409 | Positive |  |
| 28 | 160.29 | 26.816 |  |  | 78.703 | 27,460.352 | 85.947 | 23,068.291 | 87.797 | 36,958.395 | Positive | Undetermined | 6.293 | 29.730 | Positive |  |
| 29 | 160.25 | 23.713 |  |  | 78.427 | 22,485.506 | 85.356 | 16,517.178 | 87.666 | 39,520.516 | Positive | 19.563 | 20.545 | 27.191 | Positive |  |
| 30 | 159.04 | 23.325 |  |  | 78.273 | 15,894.111 | 85.407 | 22,904.027 | 87.563 | 39,710.426 | Positive | 32.910 | Undetermined | 25.874 | Positive |  |
| 31 | 159.05 | 26.059 |  |  | 78.324 | 14,713.783 | 85.459 | 21,794.189 | 87.358 | 35,491.461 | Positive | 36.402 | Undetermined | 29.195 | Positive |  |
| 32 | 160.46 | 25.074 |  |  | 78.221 | 16,500.223 | 85.407 | 18,642.883 | 87.460 | 34,221.324 | Positive | 27.702 | 28.319 | 28.798 | Positive |  |
| 33 | 159.06 | 24.499 |  |  | 78.273 | 16,530.660 | 85.613 | 22,761.973 | 87.717 | 47,821.336 | Positive | Undetermined | 5.973 | 27.723 | Positive |  |
| 34 | 159.54 | 24.670 |  |  | 78.271 | 19,032.781 | 86.271 | 21,229.328 | 88.078 | 24,151.254 | Positive | 13.584 | 17.715 | 26.804 | Positive |  |
| 35 | 159.57 | 22.130 | 82.194 | 14,912.976 |  |  | 86.220 | 25,209.344 | 88.181 | 42,081.477 | Positive | 22.510 | 27.610 | 26.111 | Positive |  |
| 36 | 160.35 | 28.021 | 81.988 | 10,891.078 | 78.323 | 15,455.877 | 86.117 | 23,845.461 | 87.871 | 25,839.080 | Positive | 23.807 | 25.584 | 31.289 | Positive |  |
| 37 | 160.38 | 24.924 | 82.142 | 37,708.629 | 78.736 | 14,730.981 | 86.065 | 23,402.859 | 87.975 | 34,013.742 | Positive | 18.380 | 18.867 | 28.032 | Positive |  |
| 38 | 162.01 | 27.500 | 82.088 | 9,153.352 | 78.535 | 15,207.086 | 85.692 | 22,415.906 | 87.545 | 31,280.473 | Positive | 34.166 | 39.534 | 30.125 | Positive |  |
| 39 | 162.02 | 26.958 | 81.985 | 27,380.613 | 78.638 | 17,346.918 | 85.846 | 27,952.066 | 87.700 | 38,928.340 | Positive | 25.260 | 29.583 | 29.976 | Positive |  |
| 40 | 162.04 | 26.188 |  |  | 78.741 | 16,577.871 | 85.795 | 28,963.525 | 87.700 | 47,975.410 | Positive | 36.757 | Undetermined | 29.645 | Positive |  |
| 41 | 166.71 | 24.747 |  |  | 78.688 | 11,270.293 | 85.927 | 21,439.764 | 87.981 | 37,717.902 | Positive | Undetermined | 4.729 | 30.542 | Positive |  |
| 42 | 163.78 | 22.556 |  |  | 78.808 | 12,128.883 | 86.207 | 28,589.445 | 88.314 | 52,690.047 | Positive | 35.586 | 36.342 | 26.186 | Positive |  |
| 43 | 166.58 | 23.713 |  |  | 78.329 | 11,961.000 | 85.722 | 24,877.676 | 87.776 | 45,893.137 | Positive | 37.791 | Undetermined | 29.400 | Positive |  |
| 44 | 164.02 | 20.956 | 81.923 | 45,010.543 | 78.483 | 12,777.211 | 85.825 | 27,201.039 | 87.776 | 44,412.332 | Positive | 17.926 | 23.174 | 26.895 | Positive |  |
| 45 | 165.97 | 24.206 |  |  | 78.756 | 11,545.430 | 85.744 | 22,294.586 | 87.954 | 37,430.465 | Positive | Undetermined | 16.919 | 28.317 | Positive |  |
| 46 | 165.51 | 26.403 | 82.250 | 35,292.453 | 78.859 | 12,163.988 | 86.104 | 20,951.469 | 87.954 | 23,271.453 | Positive | 21.657 | 24.369 | 30.709 | Positive |  |
| 47 | 166.45 | 25.670 |  |  | 77.713 | 16,580.752 | 85.825 | 23,797.518 | 87.622 | 25,014.664 | Positive | 30.117 | 32.008 | 32.034 | Positive |  |
| 48 | 166.36 | 25.809 |  |  | 78.688 | 11,241.576 | 85.825 | 21,879.023 | 87.468 | 21,958.398 | Positive | 36.458 | 38.255 | 31.007 | Positive |  |
| 49 | 165.34 | 26.686 |  |  | 78.653 | 11,652.077 | 86.053 | 21,337.355 | 87.903 | 29,255.049 | Positive | Undetermined | 14.779 | 30.160 | Positive |  |
|  |  |  |  |  |  | **All Negative** | |  |  |  |  |  |  |  |  |  |
| 50 | 157.12 | 25.568 |  |  |  |  | 85.968 | 33,745.766 | 88.029 | 48,691.023 | Negative | Undetermined | Undetermined | 26.975 | Negative |  |
| 51 | 157.39 | 25.095 |  |  |  |  |  |  | 87.823 | 35,769.059 | Negative | Undetermined | Undetermined | 25.009 | Negative |  |
| 52 | 162.07 | 24.759 |  |  |  |  |  |  | 87.648 | 37,694.355 | Negative | Undetermined | Undetermined | 29.607 | Negative |  |
| 53 | 164.12 | 23.175 |  |  |  |  |  |  | 87.878 | 32,685.543 | Negative | Undetermined | Undetermined | 28.241 | Negative |  |
| 54 | 165.07 | 22.419 |  |  |  |  | 86.207 | 22,677.789 | 88.365 | 50,947.563 | Negative | Undetermined | Undetermined | 24.317 | Negative |  |
| 55 | 163.91` | 23.279 |  |  |  |  |  |  | 88.108 | 48,142.965 | Negative | Undetermined | Undetermined | 28.96 | Negative |  |
| 56 | 165.89 | 23.832 |  |  |  |  |  |  | 88.005 | 41,674.066 | Negative | Undetermined | Undetermined | 29.704 | Negative |  |
| 57 | 164.54 | 23.415 |  |  |  |  | 85.722 | 22,421.445 | 87.673 | 42,630.070 | Negative | Undetermined | Undetermined | 30.306 | Negative |  |
|  |  |  |  |  |  | **SYBR Positive_commercial kit Negative** | | | | | |  |  |  |  |  |
|  |  |  |  |  |  |  | | | | | |  |  |  |  | PCR result |
| 58 | 160.06 | 27.309 |  |  | 78.168 | 15,420.802 | 87.768 | 24,404.719 | 89.833 | 16,689.711 | Positive | Undetermined | Undetermined | Undetermined | Negative | positive |
| 59 | 157.14 | 26.808 |  |  | 77.982 | 20,171.916 | 85.917 | 24,823.512 | 87.874 | 41,691.055 | Positive | Undetermined | Undetermined | 29.760 | Negative | negative |
| 60 | 157.21 | 26.788 | 79.528 | 23,400.113 | 77.725 | 23,190.389 | 85.968 | 17,638.922 | 87.720 | 15,171.685 | Positive | Undetermined | Undetermined | 29.545 | Negative | negative |
| 61 | 156.42 | 27.675 |  |  | 77.982 | 11,823.851 | 85.968 | 14,053.350 | 87.977 | 13,897.023 | Positive | Undetermined | Undetermined | 29.293 | Negative | positive |
| 62 | 156.94 | 25.898 | 80.971 | 10,278.659 | 77.364 | 10,502.136 | 85.762 | 18,522.346 | 87.823 | 23,491.320 | Positive | Undetermined | Undetermined | 28.947 | Negative | positive |
| 63 | 158.27 | 24.261 |  |  | 77.879 | 12,738.994 | 85.710 | 18,782.756 | 87.720 | 34,102.215 | Positive | Undetermined | Undetermined | 27.657 | Negative | undetermined |
| 64 | 100.104L | 28.038 | 81.942 | 17,533.844 | 78.653 | 23,725.438 | 85.847 | 20,334.436 | 87.851 | 33,280.203 | Positive | Undetermined | Undetermined | 31.901 | Negative | undetermined |
| 65 | 100.112L | 27.244 | 81.891 | 14,826.096 | 78.499 | 18,034.703 | 85.796 | 24,370.982 | 87.748 | 37,031.297 | Positive | Undetermined | Undetermined | 30.827 | Negative | positive |
| 66 | 100.136L | 23.441 | 82.045 | 12,308.043 | 78.551 | 16,719.590 | 85.847 | 31,664.225 | 88.057 | 53,096.879 | Positive | Undetermined | Undetermined | 25.221 | Negative | positive |
| 67 | 100.145L | 21.827 |  |  | 78.294 | 12,735.380 | 85.744 | 22,467.652 | 87.697 | 39,269.410 | Positive | Undetermined | Undetermined | 25.623 | Negative | undetermined |
| 68 | 116.025L | 24.026 | 81.513 | 9,875.732 | 78.176 | 16,113.243 | 85.415 | 22,907.361 | 87.469 | 39,798.813 | Positive | Undetermined | Undetermined | 28.457 | Negative | positive |
| 69 | 100.090L | 22.820 | 81.891 | 9,210.04 | 78.294 | 13,493.40 | 85.796 | 25,724.49 | 87.748 | 38,834.20 | Positive | Undetermined | Undetermined | 26.922 | Negative | positive |
| 70 | 159.07 | 21.407 |  |  | 78.529 | 19,364.176 | 85.715 | 23,224.367 | 87.922 | 49,715.895 | Positive | Undetermined | Undetermined | 23.940 | Negative | undetermined |
| 71 | 159.08 | 24.074 |  |  | 78.529 | 26,065.836 | 85.715 | 27,950.270 | 87.820 | 48,458.414 | Positive | Undetermined | Undetermined | 27.850 | Negative | undetermined |
| 72 | 160.31 | 27.311 |  |  | 78.703 | 19,249.354 |  |  | 87.900 | 29,258.752 | Positive | Undetermined | Undetermined | 30.526 | Negative | negative |
| 73 | 160.04 | 25.430 | 80.749 | 15,519.137 | 78.530 | 18,818.221 | 86.271 | 18,506.064 | 87.975 | 22,056.859 | Positive | Undetermined | Undetermined | 29.400 | Negative | negative |
| 74 | 160.16 | 25.177 | 82.091 | 11,628.271 | 78.581 | 15,861.441 | 86.013 | 28,269.074 | 88.078 | 56,295.973 | Positive | Undetermined | Undetermined | 30.195 | Negative | positive |
| 75 | 162.03 | 24.258 |  |  | 78.638 | 18,802.588 | 85.898 | 29,248.230 | 87.751 | 45,763.887 | Positive | Undetermined | Undetermined | 26.295 | Negative | negative |
| 76 | 162.05 | 34.130 |  |  | 77.506 | 25,844.852 |  |  | 87.339 | 9,697.884 | Positive | Undetermined | Undetermined | 22.837 | Negative | negative |
| 77 | 164.27 | 23.348 |  |  | 78.380 | 11,198.582 |  |  | 87.930 | 50,414.348 | Positive | Undetermined | Undetermined | 27.880 | Negative | negative |
| 78 | 166.61 | 23.369 |  |  | 78.586 | 10,982.369 |  |  | 87.724 | 43,584.203 | Positive | Undetermined | Undetermined | 28.188 | Negative | undetermined |
| 79 | 164.36 | 25.533 |  |  | 78.637 | 11,091.994 | 85.876 | 17,648.221 | 87.776 | 29,974.148 | Positive | Undetermined | Undetermined | 29.843 | Negative | positive (seq) |
| 80 | 164.47 | 27.410 |  |  | 78.534 | 11,815.230 | 85.876 | 22,065.617 | 87.827 | 28,747.170 | Positive | Undetermined | Undetermined | 32.212 | Negative | negative |
| 81 | 165.44 | 25.877 |  |  | 78.345 | 9,453.904 |  |  | 87.954 | 33,168.156 | Positive | Undetermined | Undetermined | 31.601 | Negative | und  termined |
| 82 | 166.24 | 21.729 | 81.820 | 8,159.522 |  |  | 85.722 | 22,657.387 | 87.878 | 44,914.289 | Positive | Undetermined | Undetermined | 27.451 | Negative | undetermined |
| 83 | 164.42 | 24.508 |  |  | 78.329 | 9,015.851 | 85.619 | 17,147.021 | 87.519 | 24,546.426 | Positive | Undetermined | Undetermined | 31.396 | Negative | negative |
| 84 | 165.27 | 26.292 | 81.685 | 9,289.496 | 78.551 | 11,500.439 | 85.899 | 24,221.045 | 87.748 | 29,584.457 | Positive | Undetermined | Undetermined | 32.030 | Negative | negative |
|  |  |  |  |  |  | **SYBR Negative_Sansure Positive** | | | | |  |  |  |  |  | PCR result |
| 85 | 165.02 | 22.897 |  |  |  |  |  |  | 87.724 | 41,292.852 | Negative | 34.876 | 35.564 | 24.949 | Positive | negative |
| 86 | 165.104 | 23.836 |  |  |  |  |  |  | 87.954 | 30,614.727 | Negative | 36.431 | Undetermined | 28.525 | Positive | positive (seq) |
| 87 | 166.03 | 25.313 |  |  |  |  | 85.773 | 25,382.316 | 87.776 | 48,181.316 | Negative | 36.670 | Undetermined | 29.601 | Positive | negative |
| 88 | 162.06 | 24.000 |  |  |  |  | 85.589 | 30,429.172 | 87.494 | 55,936.672 | Negative | 36.503 | Undetermined | 28.888 | Positive | negative |
| 89 | 162.08 | 25.144 |  |  |  |  | 85.486 | 23,657.658 | 87.597 | 43,172.418 | Negative | 35.260 | Undetermined | 29.626 | Positive | positive |
| 90 | 166.63 | 23.463 |  |  |  |  | 85.927 | 25,674.121 | 87.981 | 46,152.188 | Negative | Undetermined | 16.902 | 26.877 | Positive | negative |

**Supplementary Table 03**

| **Primer Combination** | **Sample ID** | **Ct Value** | **Melting Temperature** | **Derivative Reporter** |  |
| --- | --- | --- | --- | --- | --- |
| **Uniplex Assay** | | | | | |
| JUST_N1 | 103.49 | 28.865 | 82.117 | 49,427.047 | JUST_N1 Specific |
|  | 103.59 | 27.081 | 82.422 | 44,041.594 |  |
|  | 105.92 | 24.958 | 82.422 | 33,674.445 |  |
|  | 105.82 | 31.496 | 74.949 | 33,807.930 | Non-specific |
|  |  |  | 80.897 | 16,299.283 |  |
| JUST_E1 | 103.49 | 28.266 | 79.046 | 40,590.711 | JUST_E1 Specific |
|  | 103.59 | 26.847 | 79.504 | 50,604.645 |  |
|  | 105.92 | 25.883 | 79.656 | 48,298.816 |  |
|  | 105.82 | 33.038 | 66.095 | 10,334.008 | Non-specific |
|  |  |  | 82.246 | 20,230.045 |  |
|  |  |  | 85.141 | 17,941.426 |  |
| JUST_S1 | 103.49 | 25.997 | 76.324 | 51,881.965 | JUST_S1 Specific |
|  | 103.59 | 25.233 | 76.629 | 34,492.000 |  |
|  | 105.92 | 23.581 | 76.629 | 36,840.020 |  |
|  | 105.82 | 31.273 | 67.628 | 9,944.588 | Non-specific |
|  |  |  | 74.341 | 30,616.861 |  |
| JUST_RdRp1 | 103.49 | 27.599 | 77.366 | 50,752.266 | JUST_RdRp1 Specific |
|  | 103.59 | 26.594 | 77.671 | 50,150.043 |  |
|  | 105.92 | 24.918 | 77.671 | 41,050.215 |  |
|  | 105.82 | 34.294 | 80.413 | 28,342.781 | Non-specific |
| Beta-actin | 117.199 | 23.377 | 87.728 | 51,152.402 | Beta-actin Specific |
|  | 100.115 | 25.559 | 87.470 | 49,336.742 |  |
| GAPDH | 117.199 | 31.161 | 82.928 | 63,028.238 | GAPDH Spcific |
|  |  |  | 69.481 | 17,801.490 | Non-specific |
|  | 100.115 | 36.145 | 82.979 | 45,427.840 | GAPDH Spcific |
| **Duplex Assay** | | | | | |
| JUST_N1  +JUST_E1 | 103.59 | 25.944 | 80.899 | 18,224.209 | JUST_N1 Specific |
|  |  |  | 77.696 | 9,732.452 | JUST_E1 Specific |
|  | 117.199 | 24.647 | 82.278 | 31,206.523 | JUST_N1 Specific |
|  |  |  | 78.905 | 97,134.820 | JUST_E1 Specific |
| JUST_N1  +JUST_S1 | 103.59 | 26.613 | 81.662 | 16,774.512 | JUST_N1 Specific |
|  |  |  | 75.408 | 11,924.494 | JUST_S1 Specific |
|  | 117.199 | 24.178 | 82.278 | 16,230.109 | JUST_N1 Specific |
|  |  |  | 75.788 | 54,621.520 | JUST_S1 Specific |
| JUST_N1  +JUST_RdRp1 | 103.59 | 26.774 | 81.967 | 18,510.281 | JUST_N1 Specific |
|  |  |  | 77.086 | 19,971.842 | JUST_RdRp1 Specific |
|  | 117.199 | 22.934 | 82.227 | 25,829.594 | JUST_N1 Specific |
|  |  |  | 76.861 | 67,849.789 | JUST_RdRp1 Specific |
| JUST_E1  +JUST_S1 | 117.199 | 24.265 | 78.956 | 80,005.727 | JUST_E1 Specific |
| JUST_E1  +JUST_RdRp1 | 117.199 | 23.429 | 78.905 | 61,383.828 | JUST_E1 Specific |
|  |  |  | 76.605 | 25,566.580 | JUST_RdRp1 Specific |
| JUST_S1  +JUST_RdRp1 | 117.199 | 22.878 | 77.014 | 68,878.430 | JUST_RdRp1 Specific |
| JUST_N1  +GAPDH | 117.199 | 24.509 | 82.267 | 51,459.141 | JUST_N1 Specific |
|  |  |  | 83.905 | 29,276.434 | GAPDH Specific |
|  |  |  | 73.361 | 14,140.837 | Non-specific |
| JUST_E1  +GAPDH | 117.199 | 27.825 | 78.889 | 62,769.035 | JUST_E1 Specific |
|  |  |  | 82.421 | 23,866.477 | GAPDH Specific |
| JUST_S1  +GAPDH | 117.199 | 24.897 | 75.818 | 48,897.164 | JUST_S1 Specific |
|  |  |  | 82.421 | 14,717.176 | GAPDH Specific |
| **Triplex** | | | | | |
| JUST_N1  +JUST_E1  +JUST_S1 | 103.59 | 26.525 | 82.425 | 12,021.470 | JUST_N1 Specific |
|  |  |  | 76.018 | 6,761.184 | JUST_S1 Specific |
| JUST_N1  +JUST_RdRp1  +JUST_S1 | 103.59 | 28.707 | 82.272 | 8,179.607 | JUST_N1 Specific |
|  |  |  | 77.238 | 12,854.509 | JUST_RdRp1 Specific |
| JUST_N1  +JUST_RdRp1  +JUST_E1 | 103.59 | 26.544 | 82.272 | 10,984.571 | JUST_N1 Specific |
|  |  |  | 79.069 | 8,844.529 | JUST_E1 Specific |
| JUST_N1  +JUST_E1  +Beta-actin | 117.199 | 22.999 | 81.825 | 29,259.492 | JUST_N1 Specific |
|  |  |  | 78.487 | 28,967.324 | JUST_E1 Specific |
|  |  |  | 85.780 | 29,502.145 | Beta-actin specific |
|  |  |  | 87.731 | 42,329.117 |  |
| JUST_N1  +JUST_E1  +GADPH | 117.199 | 23.226 | 82.203 | 56,078.969 | JUST_N1 Specific |
|  |  |  | 78.434 | 15,534.940 | JUST_E1 Specific |
| **Quadruplex** | | | | | |
| JUST_N1  +JUST_E1  +JUST_S1  +JUST_RdRp1 | 103.59 | 26.805 | 82.120 | 4,701.885 | JUST_N1 Specific |
|  |  |  | 77.543 | 9,604.580 | JUST_RdRp1 Specific |

**supplementary table 4: Cost of the total process per sample**

| **Reagents and Consumables** | **cost (per sample)** |
| --- | --- |
| Lucigen RNA extraction kit | 10 BDT |
| Luna RT-PCR kit | 120 BDT |
| Primers | 3 BDT |
| Consumables | 7 BDT |
| Total | 140 BDT ($1.65) |

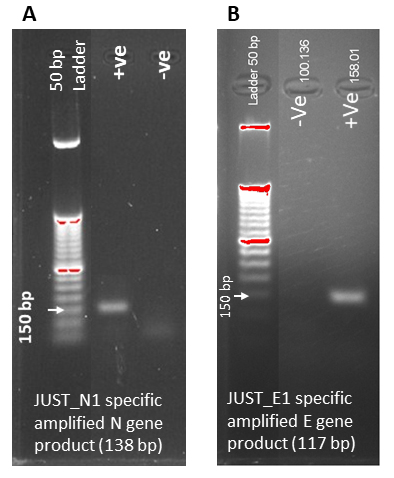

**Supplementary Figure 01: Gel electropherogram of RT-PCR amplified products**

**Supplementary Figure S2**

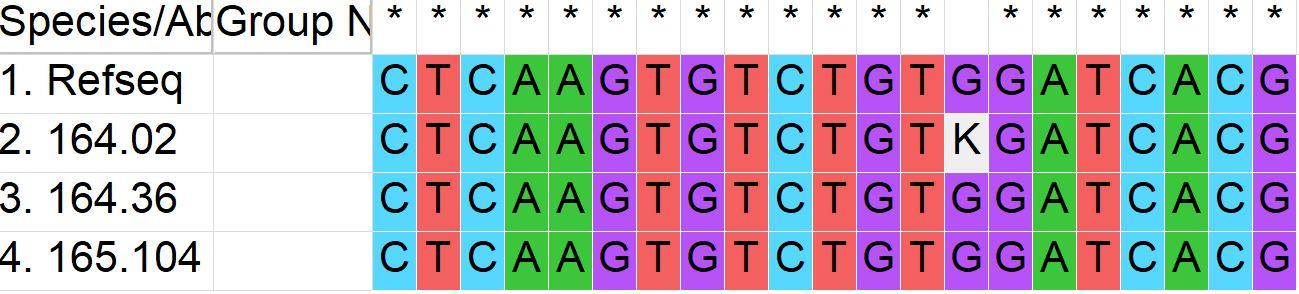
